# Improving the cost efficiency of preventive chemotherapy: impact of new diagnostics on stopping decisions for control of schistosomiasis

**DOI:** 10.1101/2023.09.25.23296064

**Authors:** Luc E. Coffeng, Matthew Graham, Raiha Browning, Klodeta Kura, Peter J. Diggle, Matthew Denwood, Graham F. Medley, Roy M Anderson, Sake J. de Vlas

**Author notes:** Corresponding author; postal address: Erasmus MC, Department of Public Health, P.O. box 2040, 3000 CA Rotterdam, The Netherlands.

## Abstract

**Background:** Control of several neglected tropical diseases (NTDs), including schistosomiasis, relies on the regular distribution of preventive chemotherapy (PC) over many years. For the sake of sustainable NTD control, a decision must be made at some stage to scale down or stop PC. These ‘stopping decisions’ are based on population surveys that assess whether infection levels are sufficiently low (typically less than 1%). For schistosomiasis control, concerns have been raised regarding the limited sensitivity of the currently-used diagnostic (Kato-Katz or KK) to detect low intensity infections. The use of new, more sensitive, molecular diagnostics has been proposed.

**Methods:** Through statistical analysis of *Schistosoma mansoni* egg counts collected from Burundi and a simulation study using an established transmission model for schistosomiasis, we investigated the extent to which more sensitive diagnostics can improve decision making regarding stopping or continuing PC for the control of *S. mansoni*.

**Results:** We found that KK-based strategies perform reasonably well for determining when to stop PC at a local scale. Use of more sensitive diagnostics only leads to a marginally improved health impact (person-years lived with heavy infection) and comes at a cost of continuing PC for longer, unless the decision threshold for stopping PC is adapted upwards. However, if this threshold is set too high, PC may be stopped prematurely, resulting in a rebound of infection levels.

**Conclusions:** We conclude that the potential value of more sensitive diagnostics lies more in the reduction of survey-related costs than in the direct health impact of improved parasite control.

**summary:** Compared to Kato-Katz faecal thick smears, model-based analyses suggest that the use of more sensitive tests only marginally changes the impact of decisions to locally stop preventive chemotherapy against schistosomiasis in terms of disease burden (person-years lived with heavy infection).

## Introduction

Neglected tropical diseases (NTDs) are a group of diseases that cause a significant health and socio-economic burden that mostly impacts the poorest parts of the world [1]. Preventive chemotherapy (PC) is a central component in the control and elimination of several NTDs, including trachoma and helminth infections like schistosomiasis [2]. PC constitutes the blanket treatment of populations in endemic areas, regardless of individuals’ infection status. With regular distribution and high enough population coverage, PC is effective at reducing infection levels and morbidity in endemic populations. PC may even lead to complete local interruption of transmission, although sustainable large-scale elimination should also be expected to require behavioural and structural interventions such as improved access to water, sanitation and hygiene [3,4]. For the sustainability of NTD control, guidelines by the World Health Organization (WHO) recommend that PC is scaled down or stopped when infection levels in target populations have decreased to sufficiently low levels. This is also the case for the disease schistosomiasis (SCH), which is caused by a parasitic helminth infection that is currently primarily controlled via school-based PC with the drug praziquantel.

SCH is transmitted via contamination of fresh water bodies with human faeces or urine and intermediate fresh-water snail hosts that releases cercarial infective stages [5]. The majority of human SCH infections are caused by two species, where the adult male-female worm pairs either reside in the venules of the intestines (*Schistosoma mansoni*) or the bladder (*S. haematobium*). In endemic areas, infection levels typically peak in school age children (SAC), although adults can also harbour a substantial fraction of the worm population [6,7]. The distribution of adult worms across the human population is highly overdispersed, meaning that a few individuals are infected with many worms (up to hundreds or thousands [8]) but most individuals carry only a few or no worms [5]. As a result, a single treatment with praziquantel, which kills ∼86% of the adult worms [9], is unlikely to eliminate infection in individuals with higher infection intensities. These individuals, along with untreated individuals, are important reservoirs of infection from which transmission continues between PC rounds. Therefore, PC must be implemented repeatedly and at sufficiently high coverage levels in order to successfully reduce infection to low levels [10].

An increasing number of SCH-endemic areas are approaching a point where, after 5 to 6 years of PC, it may be possible to scale down or even stop PC if the prevalence of infection in SAC is low enough, e.g., 2% [11]. The WHO-recommended diagnostic technique to measure infection levels is microscopy-based detection of SCH eggs by either Kato-Katz (KK) faecal smears or urine filtration. However, an often-raised concern is that these detection methods are suboptimal due to their poor sensitivity to detect low-intensity infections [12], which constitute the majority of infections, especially after repeated PC rounds. The concern is that decisions based on low-sensitivity tests may lead to prematurely stopping PC and that infection levels will quickly bounce back. Another concern is that when population infection levels are very low, the cost of a survey based on the currently recommended diagnostics may be higher (due to required high sample sizes) in some settings than the delivery of a round of PC itself [12]. It has therefore been suggested that policy decisions should be based on more sensitive new molecular tests [13].

In this study, we investigate the extent to which the use of more sensitive diagnostic techniques can contribute to improved decision-making for PC. We hypothesise that the use of better diagnostic techniques may improve the identification of populations that do and do not need PC, leading to the same or improved health impact, possibly with fewer PC treatments distributed. Here, we focus on populations that are endemic for *S. mansoni* (intestinal SCH) where PC is targeted at SAC. We employ an existing SCH transmission model [14–17] to simulate the impact of PC and the outcome of different diagnostics strategies to inform stopping decisions for PC. To accurately capture how the sensitivity of KK changes with individual worm burden, we analysed historical data representing seven days of duplicate *S. mansoni* egg counts from Burundi [18]. We then compared the performance of decision strategies based on single and duplicate KK, as well as a range of new hypothetical diagnostic tests with higher sensitivity for detection of low intensity infections.

## Methods

### Mathematical model for trends of *S. mansoni* infection during and after PC

We employed a published previously age-structured individual-based stochastic model [14– 17]. The model assumes that *S. mansoni* worms are monogamous and that their distribution has a negative binomial form. The within-host section of the model describes the evolution of the worm burden in individuals as a function of age. The model was implemented in Python, for which the code can be found here: https://github.com/NTD-Modelling-Consortium/ntd-model-sch/tree/Endgame_v2. A mathematical description of a deterministic version of the model is provided in the Supplementary Data. There, we also describe how we adhered to the PRIME-NTD principles (Policy-Relevant Items for Reporting Models in Epidemiology of Neglected Tropical Diseases) [19].

The model was expanded with mechanisms to dynamically stop PC during the simulation, conditional on a survey result being under a user-defined threshold. Further, to better capture the sensitivity of KK variants, we updated model concepts for simulation of egg counts by adding structured variation of egg counts by day and by repeated slide, which were governed by two shape parameters *k*_*day*_ and *k*_*slide*_ (lower values indicate higher variation and values *k* ≥5are suggestive of little evidence for overdispersion; for technical details, see Supplementary Data). We further added model concepts for new hypothetical diagnostic tests that can detect individual worms. To capture that the sensitivity of such tests increases with the intensity of infection, we defined sensitivity *S*_*t*_ of test *t* as a function of the number of adult worms *N* and the probability *P*_*t*_ that the test can detect a single worm. This means that 1 − *P*_*t*_ is the probability that a worm will escape detection and that (1 − *P*_*t*_)^*N*^ is the probability that all *N* worms will escape detection. Therefore, overall test sensitivity was defined as *S*_*t*_ = 1 − (1 − *P*_*t*_)^*N*^.

### Quantification of diagnostic variation in faecal egg counts based on Kato-Katz

To quantify the two shape parameters *k*_*day*_ and *k*_*slide*_ that govern variation in individuals’ faecal egg counts by day and slide, we analysed a historical dataset comprising seven days of duplicate *S. mansoni* egg counts from 200 individuals in Burundi [18]. These data were based on KK slides of 1/40 = 0.025 gram faeces, which deviates from the recommended 1/24 = 0.417 gram that is typically used nowadays. Egg counts were modelled using a Bayesian statistical model, assuming that counts follow an overdispersed Poisson distribution that captures variation between individuals, between days, and between repeated slides based on the same faecal sample [20] (for technical details, see Supplementary Data).

### Simulating the impact of different diagnostic strategies for decisions to stop PC

Simulations were run for a population of 3,000 humans, which was taken to represent a small homogenous transmission area. Transmission conditions in terms of the basic reproductive number *R*_0_ (recording the average transmission level in the population) and a shape parameter *k* (individual exposure heterogeneity) were allowed to vary randomly between repeated simulations such that the baseline prevalence of infection in SAC was between 0% and 25% (based on single KK). For each transmission condition, we simulated five years of PC targeting SAC at 90% coverage. One year after the fifth PC round, just prior to a sixth round, a survey was simulated that tested 450 SAC (about half of SAC) with one of various diagnostic strategies (details below). If the survey resulted in a prevalence estimate under the user-defined threshold, PC was automatically stopped after the sixth PC round, and if not, PC was continued and another survey was done four years later. Surveys were simulated every four years until a decision to stop PC was reached.

Decisions to stop PC were based on either single KK, duplicate KK (2 slides based on the same faecal sample), or one of six hypothetical new diagnostic tests that were characterised in terms of test sensitivity and specificity. The sensitivity of the new diagnostic tests was formulated as a per-worm-probability *P*_*t*_ to be detected. Values of *P*_*t*_ were chosen such that the lowest value corresponded to the sensitivity of duplicate KK to detect infections with 1–2 worm pairs (assuming each worm pair contributes α = 0.34 eggs to a single KK of 1/24 gram, as in the transmission model). The specificity of new diagnostic tests was assumed to be 99% or 100%; the specificity of KK was assumed to be 100%. To stop PC, we considered three thresholds for the measured infection prevalence in SAC (1%, 2%, or 5%), which were chosen for illustrative purposes as in an earlier modelling study [11]. Simulations were performed for 200 random transmission conditions, and for each of these, simulations were repeated 3 times (with different random number seeds) for a total of 600 repeated simulations per diagnostic strategy. Simulations with a baseline prevalence of 0% were discarded.

## Results

### Model quantification for sensitivity of Kato-Katz and new hypothetical diagnostic tests

Based on the Burundian KK data, we estimated that within-individual variation in egg counts by day and slide was considerable, with the shape parameters of the corresponding gamma distributions estimated at *k*_*day*_ = 1.68 (95%-Bayesian credible interval: 1.40 − 2.03) and *k*_*slide*_ = 2.37 (95%-BCI: 1.99 − 2.82). This means that variation in egg counts was driven more so by temporal sources (lower *k*) than slide-by-slide variation (higher *k*). As the data were based on 1/40 grams of faeces per KK slide, we translated our estimate of *k*_*slide*_ to a value 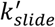 for KK slides based on 1/24 grams of faeces by setting 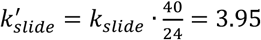. Together, *k*_*day*_ and 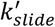 translated to a coefficient of variation of individual-level egg counts of 1.00 (95%-BCI: 0.94 − 1.07). The sensitivity of single and duplicate KK tests (based on the same faecal sample) is shown for a range of worm burdens in Figure 1.

**Figure 1.**
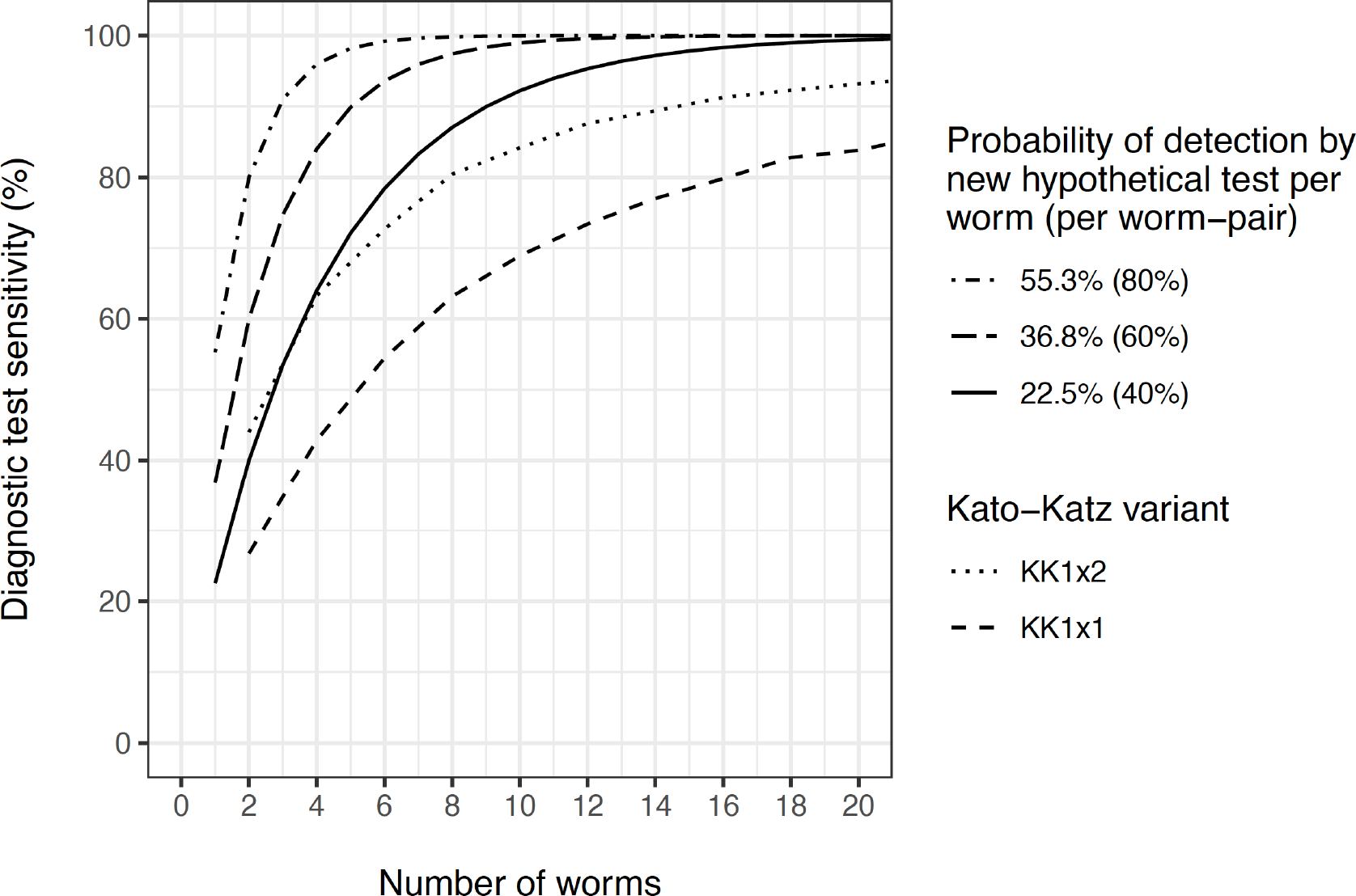
Estimated sensitivity of diagnostic tests as a function of the number of adult worms. The estimated sensitivity of Kato-Katz (KK) is conditioned on the point estimate for variation in egg counts by day (***k***_***day***_ = **1. 68**) and slide (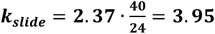, representing slides based on 1/24 gram of faeces), and the assumption that, on average, each worm pair contributes **α** = **0. 34** eggs to a single KK of 1/24 gram, as in the individual-based stochastic model. KK1x1 and KK1x2 indicate single and duplicate KK slides, respectively.

In case of 2 to 4 adult worms (or the equivalent of 1 to 2 worm pairs), the new hypothetical diagnostic test with low sensitivity (*P*_*t*_ = 22.5%, i.e., 40% per worm pair, solid line in Figure 1) performed similarly to a duplicate KK (black dotted line). Between 4 to 20 adult worms (2 to 10 worm pairs), the sensitivity of this test approached 100%, whereas the sensitivity of duplicate KK only reached around 95% for 10 worm-pairs. Therefore, for our simulations of different diagnostic strategies, we adopted three values of *P*_*t*_ for new hypothetical diagnostic tests: 22.5% (low), 36.8% (moderate), and 55.3% (high), which correspond to a 40%, 60%, and 80% probability of detection per worm pair.

### Simulating the impact of different diagnostic strategies for decisions to stop PC

We next predicted infection trends under different diagnostic strategies, based on 546 of 600 simulations for which the baseline infection prevalence was >0% (Supplementary Figure S1). If the decision to stop PC was based on KK as a diagnostic technique, the expected infection trends were very similar for single and duplicate KK (Figure 2, black lines). Further, if using a new hypothetical test with sensitivity for low worm burdens similar to KK (*P*_*t*_ = 22.5%), the predicted trends were very similar to when using KK itself (red solid lines). For a 1% decision threshold, more sensitive diagnostic tests only marginally changed expected trends (red dashed and dotted lines in left panel). For the 2% threshold, tests that were more sensitive (dashed and dotted lines) or less specific (blue lines) resulted in the decision to stop PC to be made later, on average, leading to slightly lower infection prevalence. For the 2% threshold and settings with a baseline prevalence of 5-10%, infection levels visibly rebounded, although slightly less markedly so for the more sensitive and less specific diagnostic tests. The same was the case for the 5% threshold (right panel).

**Figure 2.**
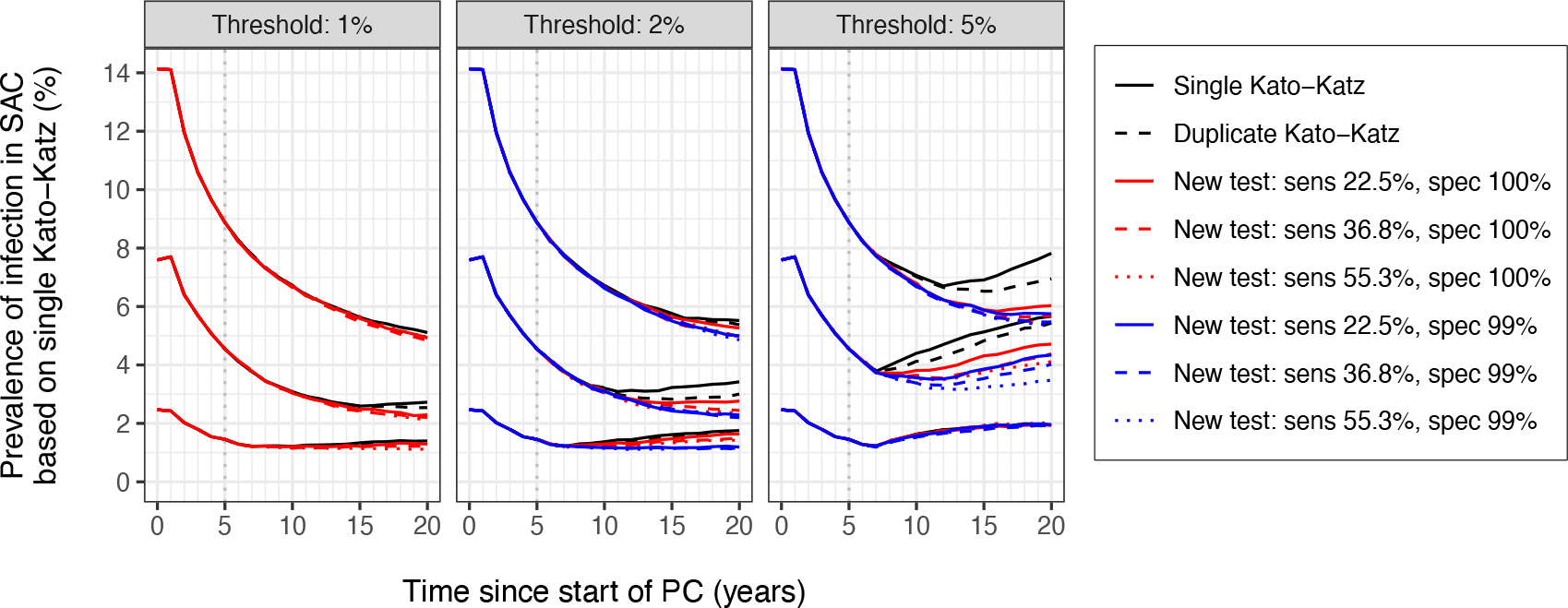
Model-predicted average trends of *Schistosoma mansoni* infection in school age children (SAC) under different diagnostic strategies for the decision to stop preventive chemotherapy. The three panels pertain to different prevalence thresholds (1%, 2%, and 5%) for making the decision to stop preventive chemotherapy. Trend lines represent averages over repeated simulations across three categories of baseline prevalence in SAC (<5%, 5-10%, and 10-25%). Note that for the scenario with 1% decision threshold (left panel), tests with 99% specificity (blue lines in other panels) were not simulated as these were considered incompatible with the threshold. Preventive chemotherapy was assumed to be implemented annually at 90% coverage of SAC.

To assess the impact of different diagnostic strategies for decisions to stop PC, we quantified the disease burden in terms of the average number of person-years with heavy intensity infections (all ages) from the start of the first survey (year 5) till 15 years later, and compared this to the average number of PC rounds that was distributed during the same period. In Figure 3, we see that for settings with a baseline prevalence of 10-15%, stopping PC based on single KK and a threshold of 2% resulted in about 9.5 person-years of heavy infection per 1,000 capita per year and distribution of on average 11.0 PC rounds. For most diagnostic strategies, a less stringent threshold of 5% resulted in fewer PC rounds (4.3 up to 11.1, depending on the diagnostic used), but came at the cost of a higher disease burden (up to 12.6 person-years of heavy infection per 1,000 capita per year). The exception was the highest sensitive test with 99% specificity which performed very similar to single KK combined with a 2% threshold. More conservative diagnostic strategies (a 1% threshold or use of more sensitive or less specific tests) led to a comparable or slightly lower disease burden, but at the cost of slightly more PC rounds. For the other two baseline prevalence categories, these patterns were qualitatively similar (Supplementary Figure 2). The probability of achieving 0% prevalence of infection within 20 years from the start of PC was 0% for settings with a baseline prevalence in SAC of ≥5%, regardless of diagnostic strategy. Only for settings with a baseline prevalence in SAC of <5%, the probability was between 4% and 6% (Supplementary Figure S3) with only small differences between diagnostic strategies.

**Figure 3.**
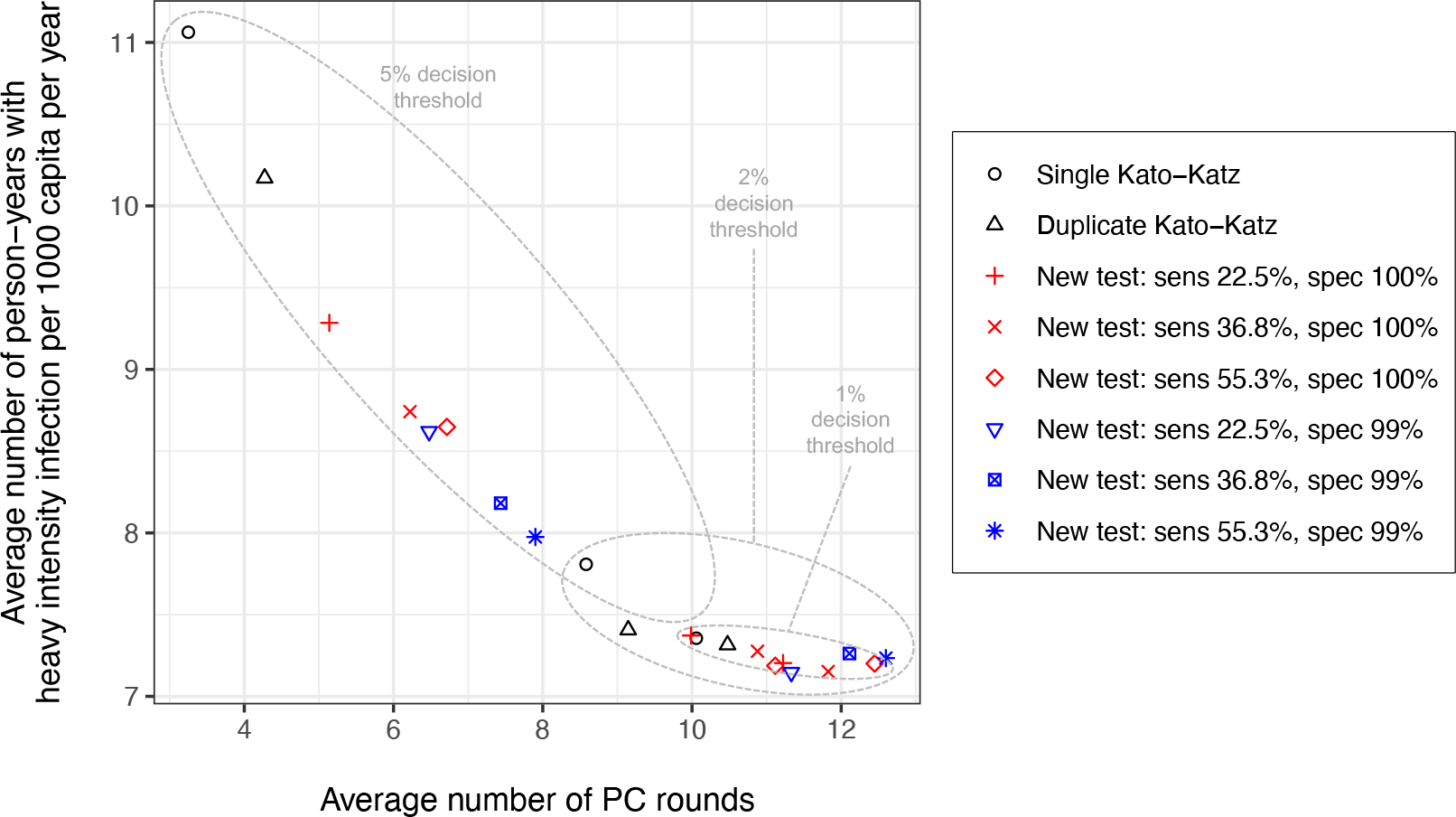
Model-predicted person-years with heavy *Schistosoma mansoni* infection in the general population versus the number of rounds of school-based preventive chemotherapy (PC) under different diagnostic strategies for the decision to stop PC. PC was assumed to be implemented annually at 90% coverage of SAC. Symbols and colors represent different diagnostic strategies to make decisions about stopping PC. Person-years with heavy infection and number of PC rounds were calculated only for the last 15 years of the simulation, i.e., from the point of the first survey at year 5 onwards and after the 6^th^ PC round had taken place. This means that at most, 14 PC rounds could have been delivered after the first survey. The result shown here represent settings with baseline prevalence of infection in school age children of 5–10%. For results in other baseline prevalence settings, see Supplementary Figure S2.

## Discussion

We illustrate that for local decisions to stop PC, the currently recommended diagnostic technique, KK, using a 2% decision threshold, almost fully minimises the disease burden (in terms of person-years with heavy infection) with the lowest possible number of PC rounds to reach that impact. New diagnostics with higher sensitivity for detection of low intensity infections may lead to a marginally lower disease burden, but at the cost of additional PC rounds. The same outcome can also be achieved by lowering the decision threshold for KK-based decisions to 1%. If decision thresholds for surveys based on new improved tests are adjusted upward (here to 5%) to account for higher test sensitivity, there is a risk of prematurely stopping PC. In the real world, this would mean that PC would have to be restarted (for which we do not consider the cost here); without, infection levels would likely bounce back, and via human mobility could even lead to reintroduction of infection in areas where transmission was previously interrupted. This aspect of reintroduction is currently not considered in existing SCH transmission models, which do not include a spatial component.

This study was inspired by the notion that more sensitivity diagnostic tests might contribute to more accurate decisions to stop or continue PC by improving the identification of areas that do and do not need PC [13]. Hypothetically, this would lead to the same or higher health impact with possibly fewer PC treatments distributed. However, if making local decisions about PC, we found that there is little to gain in terms of health impact by the use of new diagnostics. In fact, given the high efficacy of praziquantel, the health impact of PC against schistosomiasis is primarily driven by which age groups are targeted and the achieved population coverage [7,21]. As for the accuracy of decisions to stop or continue PC, more sensitive test (when combined with an appropriate decision threshold) should be expected to increase the probability of achieving elimination after stopping PC, although our simulation study was not powered enough to demonstrate this. In general, decision accuracy is mainly determined by pre-PC infection levels, the survey design (selection of study sites and age groups), the sample size (number of sites and number of persons per site), and the decision criterion for stopping PC [11,22–24]. Previous work has shown that the use of more sensitive diagnostic tests may allow for smaller sample sizes to achieve similar levels of accuracy in decision making, thus potentially saving costs [23]. A fair comparison of diagnostic tests with regard to the trade-off of cost and accuracy of policy decisions would require detailed cost data and information on the diagnostic variability of each diagnostic [25].

In this study, we simulated decisions to stop PC for relatively small areas (∼3,000 people) and did not consider that currently, decisions are made for larger areas where the distribution of SCH may be highly focal. Given this focality and the finite stock of praziquantel, it has been suggested that more focalised PC distribution and decision-making may be warranted [13]. For more decision-making, new diagnostics may well be more cost-efficient than Kato-Katz because of feasibility of implementation (e.g., a point-of-care lateral flow test) or lower cost per test. Quantification of these potential benefits would require either of two approaches. The first is to explicitly model transmission of infection and mobility of humans in larger geographically heterogeneous areas, for which initial attempts have been made for onchocerciasis [26,27], another helminth targeted with PC. The second approach would be a Monte Carlo simulation study of the cost and performance of spatial survey designs. Such a study would have to be informed by data on spatial heterogeneity in prevalences collected in an area where the average prevalence is around the decision threshold after multiple years of PC, analysed with either mixed effects models [28] or geospatial models.

In this study we did not explicitly consider costs of PC, diagnostics, or consequences of sub-optimal policy decisions. To better inform and design cost-efficient policies and decision strategies for the control of SCH, but also NTDs in general, there is an urgent need for estimates of the cost of making a “wrong decision”. What are the costs of continuing PC for too long, or the costs of stopping PC too early? The first is relatively straightforward and may be captured well enough by simply counting the cost per PC rounds or distributed treatment. The second, however, is much more challenging as from a program or societal perspective, in addition to the “saved cost” on PC rounds, one should also capture the cost of having to restart PC, which may require renewed investments. Hopefully, such investments are limited as national NTD control programs have matured over the last decade and are becoming more and more integrated across NTDs, meaning that stopping PC against one NTD may be less likely to lead to loss of expertise and infrastructure. However, this is speculation, and it is probably safer to assume that we would rather implement a few PC rounds too many than to stop PC too early.

In conclusion, we illustrate that compared to KK, the use of more sensitive tests for decisions to stop or continue PC at a local level will at best only marginally improve the health impact of PC programs against SCH, at the cost of potentially implementing additional PC rounds.However, more sensitive diagnostic tests may still help to correctly identify the last cases of infection and to improve the cost-efficiency of SCH control via lower cost per survey and improved feasibility of conducting surveys locally.

## Data Availability

All data produced in the present study are available upon reasonable request to the authors

## Acknowledgements

We gratefully acknowledge Amanda Minter for her coordination of the project with the Endgame team at the Bill & Melinda Gates Foundation.

## Authors’ contributions

LEC conceptualised and performed the statistical analyses of the Burundian data, conceptualised the transmission model components for simulation of Kato-Katz and new hypothetical tests, analysed and visualised the data, drafted the manuscript; MG performed the simulations with the transmission model, analysed and visualised the simulation output, and revised the manuscript; RB interpreted the data and revised the manuscript; KK interpreted the data and revised the manuscript; RMA interpreted the data and revised the manuscript; PJD interpreted the data, verified that the Burundian data did not contain residual temporal correlation that might not have been captured by the statistical model, and revised the manuscript; MD interpreted the data and revised the manuscript; GFM interpreted the data and revised the manuscript; SJdV interpreted the data and revised the manuscript.

## Supplementary Data

### Model for *Schistosoma mansoni* transmission

The model used to describe the worm burden of individuals of a given age and the quantity of infectious eggs in the environment was developed from the founding work of Anderson and May [29]. Importantly, we assume that the dynamical timescale of the miracidia, cercaria and snail intermediate host, i.e., the reciprocal of their typical respective life spans, are all very fast relative to the life span of the adult worms in the human host (hours and weeks compared with years for the adult worms). This allows us to collapse the equations describing the dynamics of these stages into the equation representing changes in adult worm load within the human host [15]. Briefly, the model is a stochastic individual-based implementation of the original partial differential equation (PDE) model describing the evolution of mean female worm burden as a function of age, *M*(*a, t*):

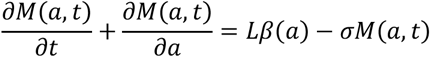

Here, *L* is the concentration of infectious material in the environment. The model describes the evolution of the female worm burden and assumes they are distributed according to an underlying negative binomial distribution. The dynamics of infectious material is governed by

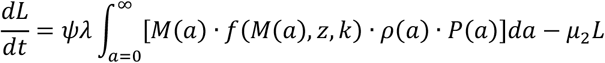

Where.*P*(*a*) is the normalised age distribution for the population. The function *f*(·) describes the production of fertile infectious material and is the product of a term representing the dampening effect of density dependent fecundity at higher worm burdens [first term] and the catalytic effect of the presence of male worms on sexual reproduction at very low worm burdens [second term] [30]:

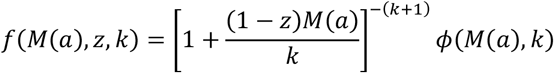

where z = *e*^−γ^ represents the strength of density-dependent fecundity and *k* is the negative binomial aggregation parameter, as discussed in the main text. The function ϕ(·) approximates the effect of monogamous sexual preproduction on egg production, where

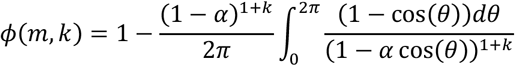

where α = *m*/(*k* + *m*). The parameter ψ characterises the flow of infectious material into the environment. This parameter and the absolute magnitude of β and ρ are subsumed into the definition of the basic reproduction number, *R*_0_, which measures the intensity of the transmission cycle.

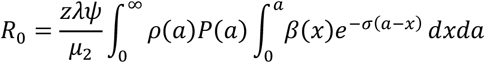

Assuming a 1:1 sex ratio in worms, the total worm burden is given by 2*M*(*a, t*). Egg counts for individual hosts of age *a, E*(*a*), can be seen as a component of the contribution of host egg output into the environment

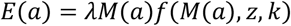

where λ is the female worm fecundity parameter.

### Updated model concepts for diagnostic test sensitivity

We employed an individual-based stochastic model for *Schistosoma mansoni* transmission that was developed and published in previous work [14–17]. Originally, in this model, egg counts based on Kato-Katz (KK) are simulated using a negative binomial draw with mean equal to the expected egg count and a single shape parameter governing the level of overdispersion between repeated egg counts. However, as soon as more than one egg count per person and time point is simulated, this approach assumes that repeated egg counts are identically independently distributed, regardless of whether they are based on the same stool sample or not. In contrast, egg counts based on repeated slides prepared from the same stool sample should be expected to be more correlated than repeated counts based on stool samples from different days.

To better capture the sensitivity of KK variants (single vs. duplicate slides based on the same faecal sample) at low levels of infection, we updated the previously used model concepts for simulation of egg counts with structured variation by day and by repeated slide. This was achieved by simulating egg counts from a gamma-gamma-Poisson compound distribution [20], where the two nested gamma distributions capture variation by day and by slide. For each simulated individual *i*, the expectation (mean) of the gamma-gamma-Poisson compound distribution was defined as in the original model [14–17] using a density-dependent function *f*(·) of the individual’s worm-pair count 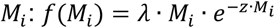, which captures that with increasing numbers of worm pairs, the number of eggs produced per worm pair decreases. Here, λ = 0.34 is the expected number of eggs per worm pair per Kato-Katz slide in absence of density dependent fecundity; and z = 0.0007 governs the degree of density dependence [17]. The degree of variation by day and slide was governed by the two shape parameters *k*_*day*_ and *k*_*slide*_ of the two nested gamma distributions, for which values were based on a statistical analysis of field data from Burundi (details in the next section below).

For simulation of surveys based on hypothetical new diagnostic tests, we developed and implemented new model concepts for detection of individual worms. Because the sensitivity of helminth diagnostics typically depends on intensity of infection, we defined sensitivity *S*_*t*_ of test *t* as a function of the number of adult worms *N* and the probability *P*_*t*_ that the test can detect a single worm. This means that the 1 − *P*_*t*_ is the probability that a worm will escape detection and that (1 − *P*_*t*_)^*N*^ is the probability that all *N* worms will escape detection. Therefore, we define overall test sensitivity as *S*_*t*_ = 1 − (1 − *P*_*t*_)^*N*^.

### Quantification of diagnostic variation in faecal egg counts based on Kato-Katz

To quantify the two shape parameters *k*_*day*_ and *k*_*slide*_ that govern variation in individuals’ faecal egg counts by day and KK slide, we analysed a historical dataset comprising seven days of duplicate *S. mansoni* egg counts from 200 individuals in Burundi [18]. We note that these data were based on KK slides of 1/40 = 0.025 gram faeces, which deviates from the recommended 1/24 = 0.417 gram that is now more typically used (this was accounted for in our analysis). Egg counts were modelled using a Bayesian statistical model, assuming that counts follow an overdispersed Poisson distribution that captures variation between individuals, between days, and between repeated slides based on the same faecal sample. These three variance components (individuals, faecal samples, and slides) were modelled using three compounded gamma distributions (parameterised in terms of shape and rate) and Poisson variation representing the counting variation in the observed egg counts [20]:

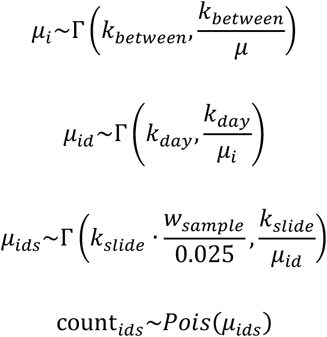

Here, *μ* represents the average egg count at the population level; *μ*_*i*_ and *μ*_*id*_ represent the expected egg count for an individual on any day and one particular day, respectively; *μ*_*ids*_ indicates the expected egg count for a single KK for a slide based on *w*_*sample*_ grams of faeces. Higher values of *k* indicate a lower coefficient of variation and therefore less overdispersion. We note that a compound gamma-Poisson distribution has the same distribution function as the negative binomial distribution, such that effectively: 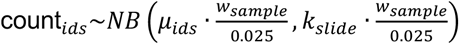, which clearly illustrates how the mean and level of overdispersion of repeated slides based on the same faecal sample change with the amount of faeces used per slide. We further note that while the mean and variance of compound gamma-gamma distributions are simple to calculate [31], the resulting distribution is not itself identical to a gamma distribution, so the overall distribution of egg counts we simulate is not exactly negative binomial.

The contribution of each variance component towards the total variation in egg counts was expressed in terms of the coefficient of variation per component (*CV*_*between*_, *CV*_*day*_, *CV*_*slide*_). For a gamma distribution with shape parameter *k*, this is defined as *CV* = *k*^− 1/2^ (and conversely, *k* = *CV*^−2^). The total coefficient of variation over all three levels is 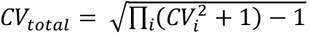. Exploratory analyses showed that specifying marginal prior distributions for individual *CV* components led to unreasonably thick tails on the push-forward prior for the total coefficient of variation *CV*_*total*_, which did not match prior expectations or the data itself. Therefore, we formulated a prior for *CV*_*total*_ and a prior for the relative contribution κ_*i*_ of each individual variance component to *CV*_*total*_, such that: 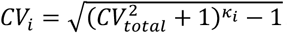. Here, we satisfy the required condition 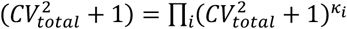 by defining κ_1:3_ as a simplex vector (i.e., ∑_*i*_ κ_*i*_ = 1, where 0 ≤ κ_*i*_ ≤ 1). For *CV*_*total*_, we defined a weakly informative half-normal prior distribution *N*^+^(0, σ_*CV0*_), with its standard deviation set to twice the empirical CV of the data (σ_*CV0*_ = 5.79). For κ_1:3_ we specified a Dirichlet prior with shape 1 for all elements (i.e., *Dir*(α_1:3_), where α_*i*_ = 1), which is a uniform prior over the simplex parameter space. For the population average egg count, we defined a weakly informative half-normal prior *N*^+^(0, σ_μ0_) with its standard deviation set to twice the empirical mean of the data (σ_μ0_ = 11.3).

The model was implemented in Stan [32], and verified by simulating data from the model, and then ensuring that we were able to recover the original input parameters by analysing the simulated data. In addition, the model was also independently implemented and tested in separate software by another author using JAGS [33] in order to further verify the inference. The final analysis was done using the Stan model in R [34] via the package *rstan* [32], using 4 parallel Markov chains and 4000 samples per chain, of which the first 2000 were discarded. In sensitivity analyses, we verified that analysis of data points from only the first three time points (days 1, 3, and 5, so excluding days, 8, 10, 32, and 37) led to the same results as when including data from all days.

## PRIME-NTD table: Policy-Relevant Items for Reporting Models in Epidemiology of Neglected Tropical Diseases

| Principle | What has been done to satisfy the principle? | Where in the manuscript is this described? |
| --- | --- | --- |
| Stakeholder engagement | The work was initiated following a request from the Bill & Melinda Gates Foundation (BMGF), Seattle. BMGF was consulted twice to clarify the research question and translate it to a model-answerable question. In a third meeting, results were presented to BMGF, and it was concluded that further research is warranted, particularly on the impact of moving from district-level to more local decision-making for PC. Further, this work has been presented at the Annual Meeting of NTD Modelling Consortium (NTDMC) held at Oxford in April, 2023, where representatives from the World Health Organization (WHO) and BMGF were present. | In this table |
| Complete model documentation | We used a previously published model, the details of which are available elsewhere (relevant sources cited in the methods section). Further, we have provided the full description of newly added model components. We further provide a link to an online repository with the Python version of the model. | Methods section and Supplementary Data |
| Complete description of data used | The data relevant for this work were quantified and described in detail in previous publications. These articles are mentioned in the Methods. | Methods section |
| Communicating uncertainty | We have considered parameter in terms of transmission conditions and a range of scenarios for performance of new diagnostics | Methods and Results sections |
| Testable model outcomes | All the model predictions are testable, provided the relevant data are collected (although this may be challenging). |  |

### Results of individual simulations

For each diagnostic strategy we performed 600 repeated simulations, which were based on 200 different parameter sets for transmission conditions and 3 repeated simulations per parameter set. This was done by first running one 5-year-long simulation for each of the 200 transmission conditions and saving the final state of the population. Then, for each diagnostic strategy, we ran 15 years of dynamic PC policy (as described in the methods in the main text), reusing the same 200 saved population states as the initial state. These last 15 years were simulated in triplicate with different random number seeds, leading to a total of 600 simulated infection trajectories per diagnostic strategy.

**Supplementary Figure S1.**
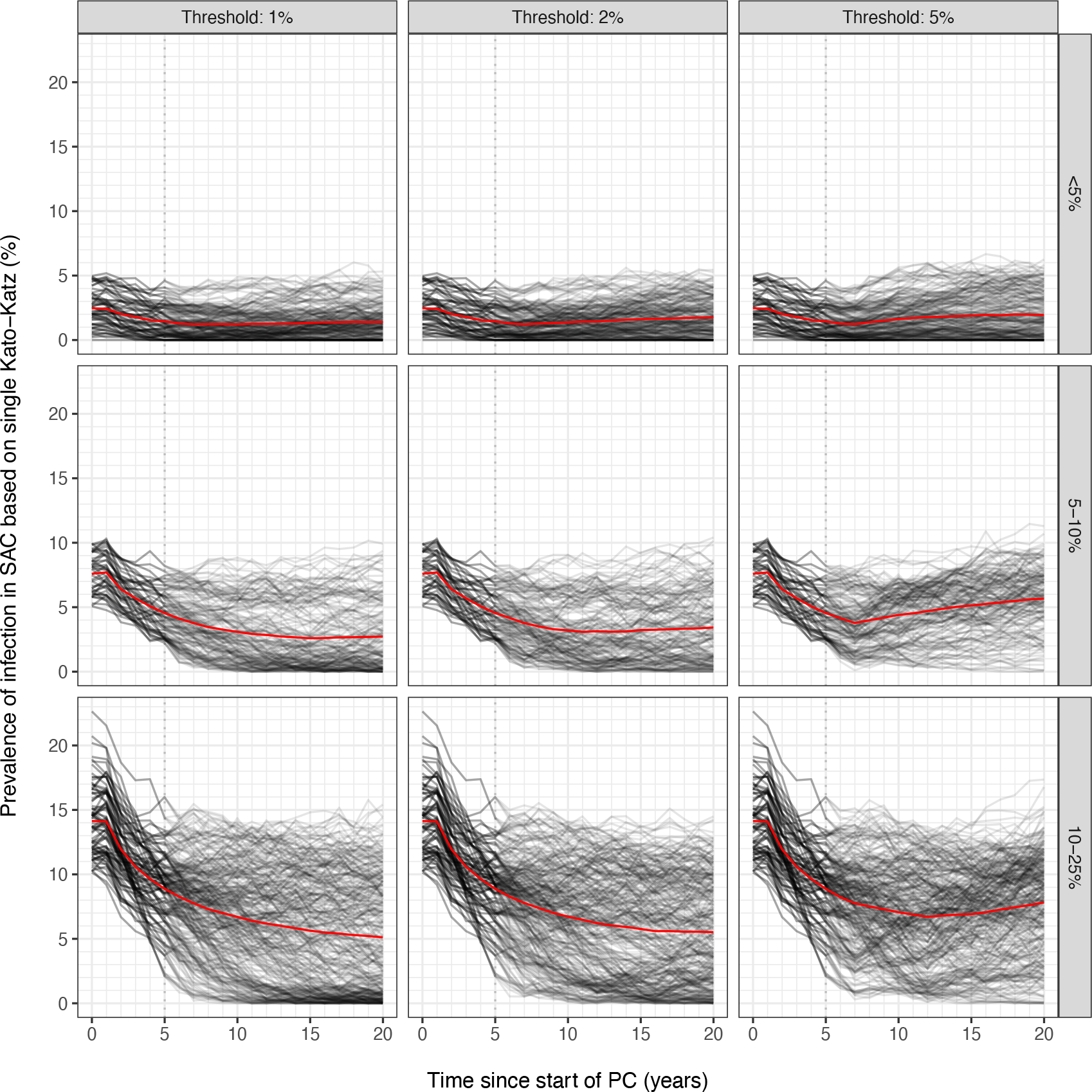
Example of model-predicted trends of *Schistosoma mansoni* infection in school age children (SAC) when using single Kato-Katz for the decision to stop preventive chemotherapy. Black lines represent individual simulation trajectories, based on simulated surveys that took place just before each PC round. Red lines represent averages over repeated simulations. The three columns of panels pertain to different prevalence thresholds (1%, 2%, and 5%) for making the decision to stop preventive chemotherapy. The three rows of panels pertain to categories of baseline prevalence of infection in SAC (<5%, 5-10%, and 10-25%). Similar type of model predictions were produced for each of the other 7 diagnostic strategies for decisions to stop PC.

Of the 600 simulations, 546 resulted in baseline prevalences of at least 0%. Based on the baseline prevalence (i.e., prevalence at time zero), these 546 simulations were categorised into three bins: <5% (N = 171), 5-10% (N = 138), and 10-25% (N = 237). Supplementary Figure S1 provides an illustration of the model-predicted infection levels, based on a decision strategy using single Kato-Katz.

For each diagnostic strategy, decision threshold, and endemicity category, we calculated the number of person-years with heavy infection (epg ≥ 400, as measured by single KK) per 1000 capita per year and the average number of rounds PC that had been distributed (Supplementary Figure S2). As the number of PC rounds and trends in infection during the first 5 simulation years were the same for all scenarios (i.e., the first survey moment), these quantities were only calculated for the last 15 simulation years only (i.e., after the first survey moment).

**Supplementary Figure S2.**
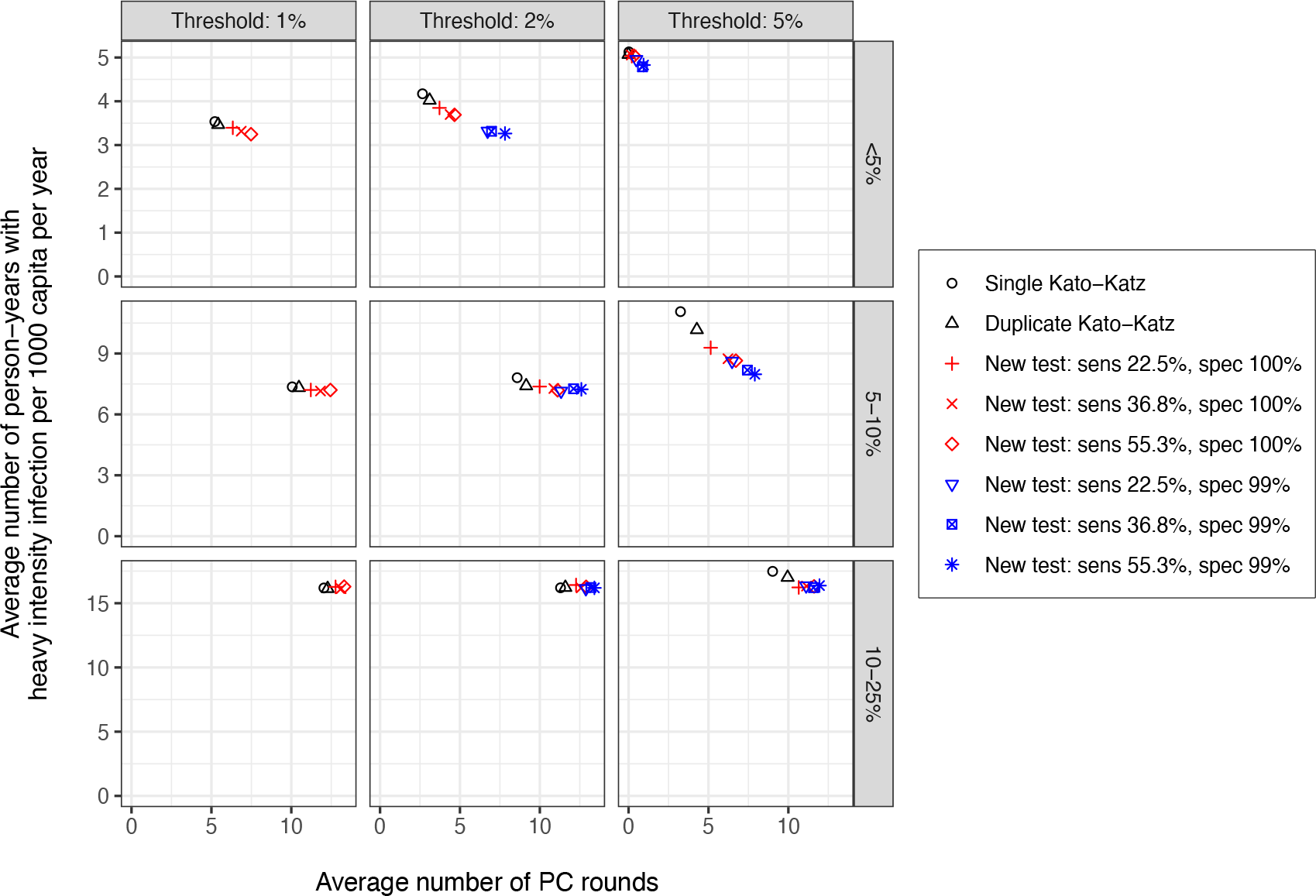
Model-predicted number of person-years with heavy *Schistosoma mansoni* infection in the general population (per 1000 capita per year) versus the average number of rounds of preventive chemotherapy (PC) targeting school age children. Symbols and colors represent different diagnostic strategies to make decisions about stopping PC. Person-years with heavy infection and number of PC rounds were calculated only for the last 15 years of the simulation, i.e., from the point of the first survey at year 5 onwards and after the 6^th^ PC round had taken place. This means that at most, 14 PC rounds could have been delivered after the first survey. The three columns of panels pertain to different prevalence thresholds (1%, 2%, and 5%) for making the decision to stop preventive chemotherapy. The three rows of panels pertain to categories of baseline prevalence of infection in SAC (<5%, 5-10%, and 10-25%).

**Supplementary Figure S3.**
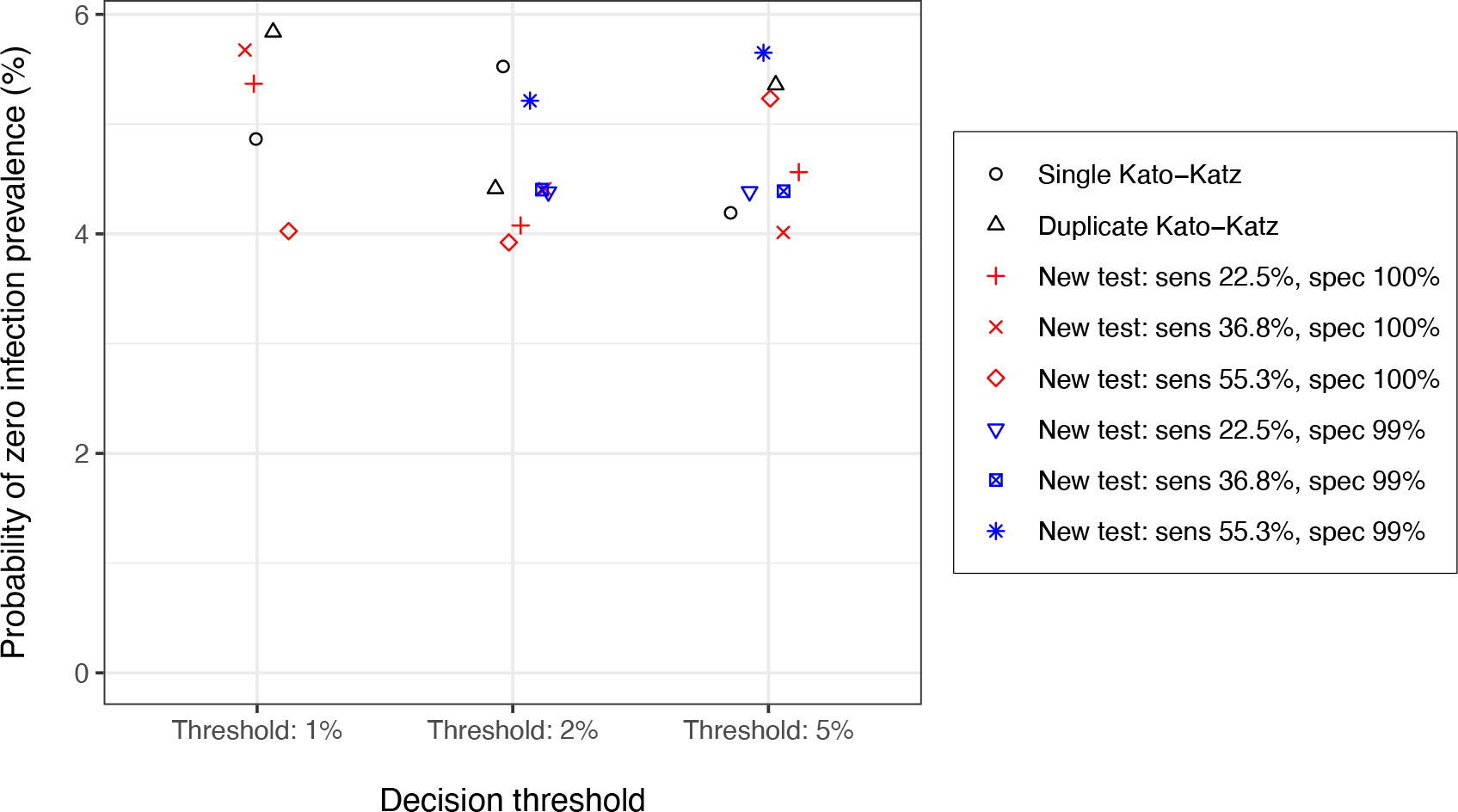
Model-predicted probability of zero infection prevalence, 20 years after the start of preventive chemotherapy (PC) targeting school age children. Symbols and colors represent different diagnostic strategies to make decisions about stopping PC. However, for all diagnostic strategies, the probability of achieving zero infection prevalence was based simulated egg counts as measured by single Kato-Katz. Results represent a setting with baseline prevalence of infection in SAC <5% where PC was implemented at 60% coverage; for the other (higher) endemicity categories, 0% infection prevalence was never achieved with PC at 60% coverage. Probabilities were based on 171 simulations, and as such, none of the probabilities differed significantly from one another.

